# A scalable platform for exon-skipping antisense oligonucleotide therapy development for inborn genetic diseases

**DOI:** 10.64898/2026.07.29.26359175

**Authors:** Logan Newton, Bushra Haque, David Cheerie, Chung Ting Tsoi, Conor Klamann, Rebecca Sakaki, Thomas Qu, Lauren Verhaeghe, Yijing Liang, Ryan M. Marks, Evgueni A. Ivakine, Ashish R. Deshwar, Gregory Costain

**Author notes:** **Address correspondence to**: Ashish Deshwar,; Gregory Costain. Equal contributions (Ashish R. Deshwar, Gregory Costain).

## Abstract

Antisense oligonucleotides (ASOs) are a versatile therapeutic modality for inborn genetic diseases. ASOs can induce skipping of “dispensable” exons containing disease-causing variants to rescue protein amount and function, but this approach has been studied for only a small number of genes. We developed a high-throughput *in silico* tool for assessing exon dispensability and designing exon-skipping ASO sequences. Parameters were optimized using known dispensable and in-frame indispensable exons. Across 72,644 exons of 5,057 disease genes, we identified thousands of new targets for exon-skipping ASOs (3.4% of exons with most stringent filters, 24.5% with less stringent filters) that collectively include 0.97%-15.6% of disease-causing variants in large-scale databases. To facilitate recognition of DNA variants potentially amenable to exon skipping as a therapeutic strategy, we established the HAWK-EYE database as a repository of exon dispensability predictions and corresponding *in silico*-optimized ASO sequences, available as an open-access web application (https://hawk-eye.research.sickkids.ca/). To illustrate translational utility, we experimentally validated a subset of the *in silico*-optimized ASO sequences that were generated for all Dispensable exons in the HAWK-EYE database, and showed that skipping a Dispensable exon in *SOX5* preserves protein function using *in vivo* and *in vitro* assays. This scalable platform approach to exon-skipping ASOs will accelerate identification and testing of amenable genetic variants.

## INTRODUCTION

Inborn genetic diseases are individually rare but collectively common, and number in the thousands^1,2^. Advances in genome diagnostics have outpaced the development and availability of disease-modifying therapies^3,4^. This unmet therapeutic need is linked to high morbidity, health-care utilization and costs, and mortality^5^. Proof-of-concept exists for individualized genetic interventions tailored to specific genetic variants and/or ultra-rare conditions^6–9^. However, current approaches to bespoke therapy development are reactive and slow^10,11^. One strategy to address this has been the development of “platform technologies” as standardized, therapeutic systems that can be readily customized for specific DNA variants^12^. Downstream regulatory reform efforts related to platform technology designations, however, must be paired with upstream efforts to accelerate amenable patient identification and preclinical experiments.

Antisense oligonucleotides (ASOs) are short, synthetic nucleotide sequences designed to selectively bind and modulate a transcript ^13^. For example, ASOs targeting exonic splicing enhancers or splice sites can prevent the recruitment of splicing machinery to an exon and result in its exclusion from a mature mRNA transcript^14,15^. Mechanisms of action are determined by chemical modifications and sequence composition^16^. Recent successes in clinical translation reflect decades of investment in fundamental and clinical research. Over 200 ASOs are currently in the development pipeline, with a modest number (<20) approved by North American and/or European regulatory bodies^17^. The N=1 Collaborative (N1C) VARIANT guidelines standardize first steps in the complex process of adjudicating DNA variants for eligibility to existing ASO mechanisms of action, including exon skipping via splice-switching^18^.

ASO-mediated exon skipping was initially pioneered as a therapeutic strategy for Duchenne muscular dystrophy^19,20^. Frame-shifting intragenic deletions and duplications in the *DMD* gene are a common cause of this progressive neuromuscular disease^19^. For select variants in this class (e.g., exon 50 deletions), excluding a neighbouring out-of-frame exon from the *DMD* transcript with an ASO (e.g., exon 51via Eteplirsen) will restore the reading frame and result in the generation of a truncated but partially functional “Becker-like” dystrophin^21^. A distinct therapeutic approach that could apply to more genes and variants involves skipping an in-frame “dispensable” exon, considered non-essential for the function or stability of the resulting protein, that harbours a pathogenic single-nucleotide variant or small insertion/deletion (indels) to rescue function^15^. The generalizability of this latter approach - beyond a small number of published examples ^22–37^ - is unknown and the genome-wide landscape of dispensable exons is uncharted^21–36^.

We developed a high-throughput pipeline for assessing the potential for functional protein product after single exon skipping, and discovered that a meaningful minority of all disease gene exons may be Dispensable (a classification of our approach). We generated optimized *in silico* ASO sequences to induce skipping of each identified Dispensable exon, and we functionally validated exon classifications and ASO-mediated exon skipping for a rare neurogenetic condition as proof-of-concept. To enable broad access to these data and support downstream therapy development, we established an open-access HAWK-EYE database that supports: (i) variant-, exon-, and gene-level queries for Dispensable exons, (ii) access to ASO sequences to support early-phase preclinical studies on patient-derived cell models, and (iii) integration of computational and experimental data.

## RESULTS

### High-throughput pipeline identifies thousands of Dispensable exons in human disease genes

To systematically assess the generalizability of single, in-frame exon skipping as a therapeutic strategy, we applied our novel pipeline (Figure 1A) to 5,057 protein-coding genes associated with human monogenic diseases (the “Mendeliome”). The resulting exon classifications were compiled into the HAWK-EYE database, establishing a comprehensive repository of exon dispensability across the Mendeliome. The 72,644 total exons in MANE Select transcripts were each classified in the HAWK-EYE database based on position and size, frame context, and predicted functional essentiality as (i) Dispensable, (ii) Indispensable, or (iii) Indeterminate (Figure 1B). Functional essentiality was inferred from exon contributions to annotated or predicted functional domains (including whether domains are uniquely encoded or redundant throughout the protein), the presence of functional elements such as motifs or post-translational modification sites, and pathogenic missense variation (see Methods). Overall, 3.4% of exons (2,481 exons across 940 unique disease genes) were classified as Dispensable (Figure 1C), with exon skipping expected to be tolerated based on stringent *in silico* filters (see Methods). Most exons were classified as Indispensable (68.4%; Figure 1C), with exon skipping expected to result in deficient protein amount and/or function. For example, 13.9% of all exons were the first or last coding exon, and an additional 50.5% were out-of-frame. The Indeterminate group (28.2% of all exons) included three tiers of increasingly strict size and functional element annotation filters (see Methods): Unlikely Dispensable (7.0% of total; 24.9% of Indeterminate), Possibly Dispensable (17.3% of total; 61.5% of Indeterminate), and Probably Dispensable (3.8% of total; 13.6% of Indeterminate) (Figure 1C). All classifications are available through the streamlined searchable HAWK-EYE database (https://hawk-eye.research.sickkids.ca/), which enables systematic interrogation of exon dispensability across genes and variants.

**Figure 1.**
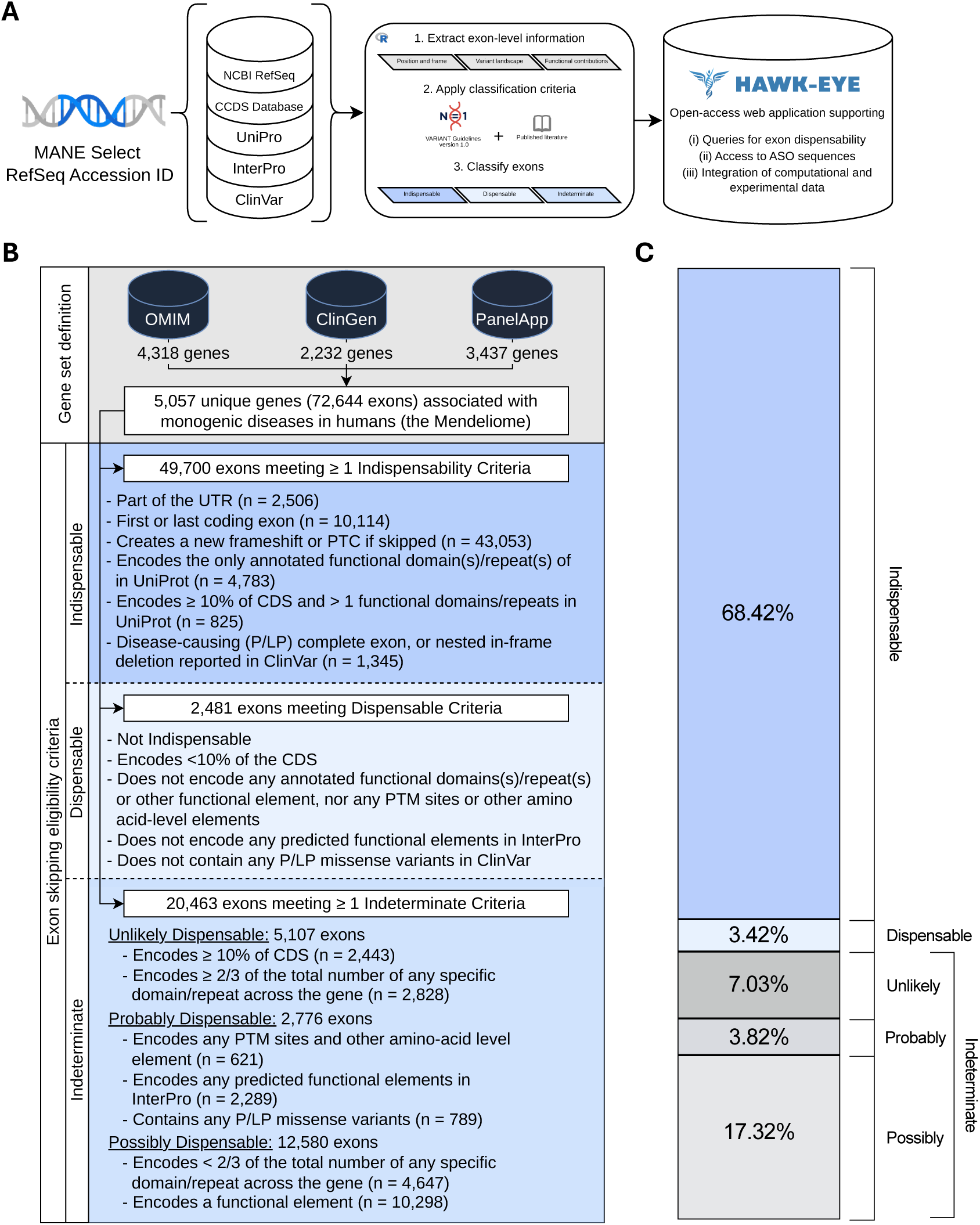
Development and Mendeliome-wide application of a high-throughput method for assessing exon (in)dispensability. (A) Using RefSeq accession IDs, exon-level annotations are aggregated from RefSeq, CCDS, UniProt, InterPro, and ClinVar. Exon features are evaluated using criteria informed by the N1C VARIANT guidelines version 1.0 and published literature to classify exons as one of Indispensable, Dispensable, or Indeterminate. Results are made available through the HAWK-EYE web application. (B) A total of 5,057 unique genes encompassing 72,644 exons were assessed by our approach. Indeterminate exons are stratified into Unlikely, Possibly, and Probably Dispensable. The number of exons satisfying individual criteria is shown for each category. Since exons are able to satisfy multiple criteria, sums of each criteria within a class may exceed the total number of exons. (C) Proportion of exon classifications across the Mendeliome. Abbreviations: UTR, untranslated region; PTC, premature termination codon; CDS, coding sequence, P/LP, pathogenic/likely pathogenic.

### Dispensable exons are characterized by distinct biological features and clinical relevance

Exon inclusion levels across all human expressed transcripts for the respective gene (median 15 transcripts per gene), averaged nucleotide-level evolutionary conservation (PhyloP) across the exon, and percent GC content are all correlated with exon functional importance and were not used to determine HAWK-EYE classifications^38–42^. The Dispensable group was significantly different from the Indispensable group, showing enrichment for exons in the bottom 20th percentile for exon inclusion, conservation, and GC content in isolation and for every combination (Figure 2A,B). Results for the Indeterminate group were intermediate between Dispensable and Indispensable (Figure 2A).

**Figure 2.**
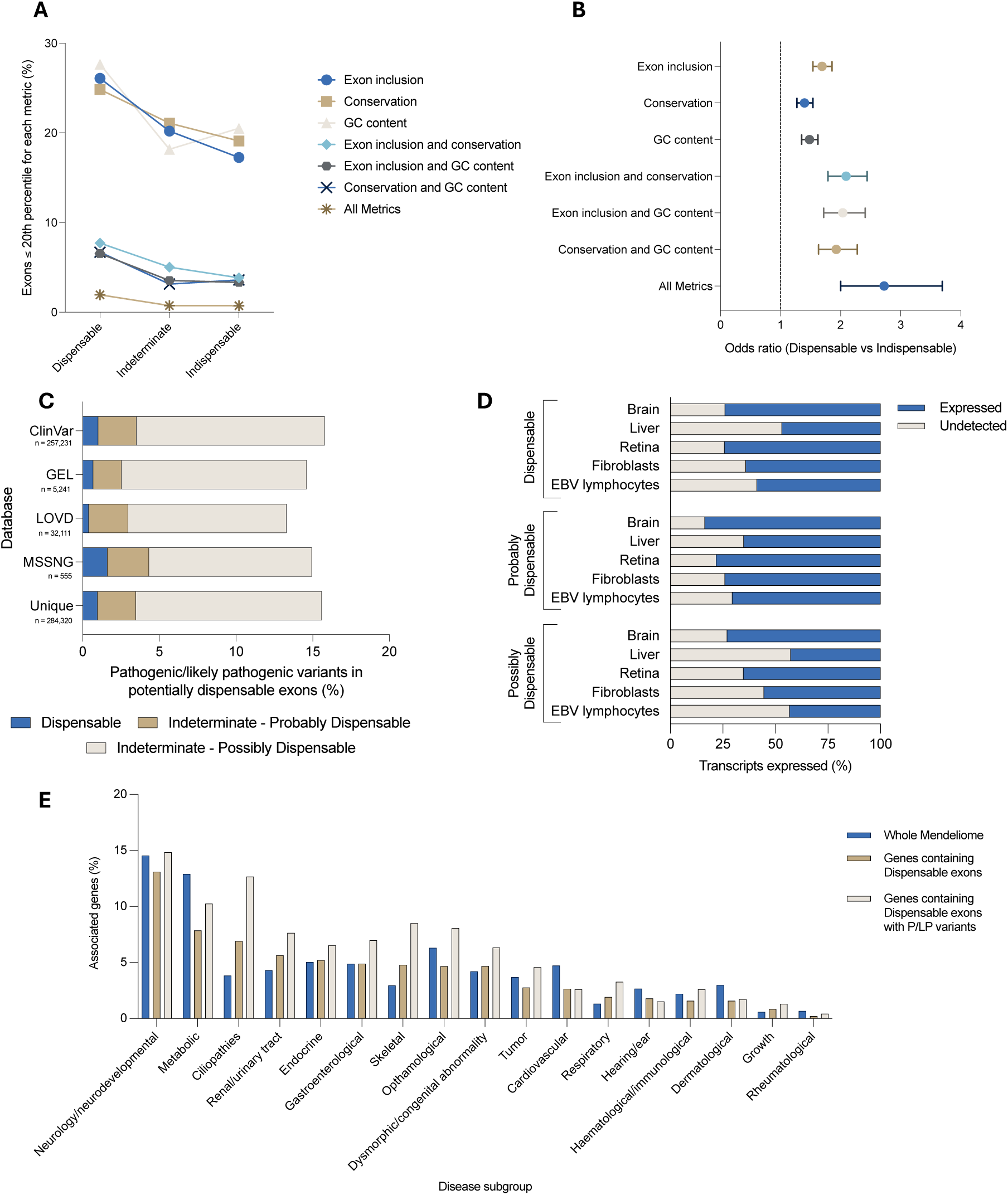
Characterization of exon dispensability predictions and associated genes. (A) Proportion of exons in each classification (Dispensable, Indeterminate, Indispensable) that exist within the 20^th^ percentile for metrics of functional importance (exon inclusion, conservation, GC content). Proportions for each metric are shown individually and in combination. (B) Odds ratio (with 95% confidence intervals) quantifying enrichment of exons within the 20^th^ percentile for each metric in the Dispensable class compared to Indispensable. (C) Proportion of pathogenic/likely pathogenic (P/LP) variants across multiple variant databases that reside in potentially dispensable exons. Variants are stratified by exon classifications. The distribution of uniquely mapped variants across all included databases is also shown. (D) Expression of MANE Select transcripts containing potentially dispensable exons that are expressed across tissues relevant to antisense oligonucleotide (ASO) delivery (brain, liver, retina) and commonly used in patient-derived cell models (fibroblasts and EBV-transformed lymphocytes). A Transcripts Per Million (TPM) threshold of 0.5 was used to define expressed/undetected transcript. (E) Percentage of genes containing Dispensable exons, genes with pathogenic/likely pathogenic (P/LP) variants in Dispensable exons, and all genes in the Mendeliome that are associated with relevant disease subgroups in the Genomics England Panel App. Genes can be associated with more than 1 disease subgroup. Figures generated using Prism GraphPad.

Across 284,320 unique disease-associated variants (pathogenic/likely pathogenic, P/LP) extracted from public and restricted-access databases, 0.97% were within exons classified as Dispensable (Figure 2C). An additional 2.5% and 12.1% of variants were within exons classified as Indeterminate – Probably Dispensable and Indeterminate – Possibly Dispensable, respectively (Figure 2C). Overall, Dispensable exons were most often in genes causing neurological/neurodevelopmental disorders, metabolic diseases, and ciliopathies (Figure 2E), where there is a precedent for ASO therapy programs. Results were similar when restricting to the subset of Dispensable exons containing known disease-associated variants (Figure 2E). Tissue-specific expression analysis on MANE Select transcripts showed that Dispensable exons are frequently expressed in tissues to which ASOs can be effectively delivered (73.9% in brain, 46.9% in liver, 74.2% in retina)^43^, and in commonly available patient-derived cell lines that facilitate early-stage pre-clinical ASO development (63.9% in fibroblasts and 58.7% in EBV-transformed lymphocytes) (Figure 2D).

### In silico-optimized Dispensable exon skipping ASOs can be designed at scale

We generated 20-mer ASO sequences to induce skipping of each Dispensable exon, based on established design principles including sequence composition, thermodynamic properties, and binding characteristics (Figure 3A). ASO sequences meeting the criteria based on these principles were generated for 2,308 of 2,481 (93.0%) Dispensable exons. We were unable to generate ASO sequences for 173 (7%) Dispensable exons primarily due to unfavourable GC content of potential ASOs, restrictive mRNA secondary structure, or high sequence homology of the target exon to other regions in the Mendeliome resulting in predicted off-target effects. We experimentally validated n=3 ASOs for each of three Dispensable exons: NM_001378454.1(*ALMS1*) exon 18, NM_001282860.2(*GON4L*) exon 6, and NM_015378.4(*VPS13D*) exon 42. Using RNA extracted from healthy control fibroblasts (GM05659) treated with ASOs, RT-PCR followed by gel electrophoresis revealed shorter amplicons consistent in size with skipping of target exons for 8 of 9 tested ASOs (Figure 3B). Exon-skipping efficiencies for these 8 ASOs were semi-quantified by densitometric analysis of band intensities, comparing residual full-length amplicons to skipped amplicons. Skipping efficiencies ranged from 21-98%, with 6 ASOs achieving >50% skipping (Figure 3B). To obtain a more precise quantification of skipping, ASOs targeting exon 18 of NM_001378454.1(*ALMS1*) were assessed by short-read bulk RNA sequencing. Exon skipping efficiencies were calculated using splice-junction reads (see Methods), revealing efficiencies of 4%, 38%, and 36% for the three ASOs (Figure 3C). A median of 12 ASO sequences per exon (interquartile range 7-19) is available for each Dispensable exon for download through the HAWK-EYE web application (https://hawk-eye.research.sickkids.ca/).

**Figure 3.**
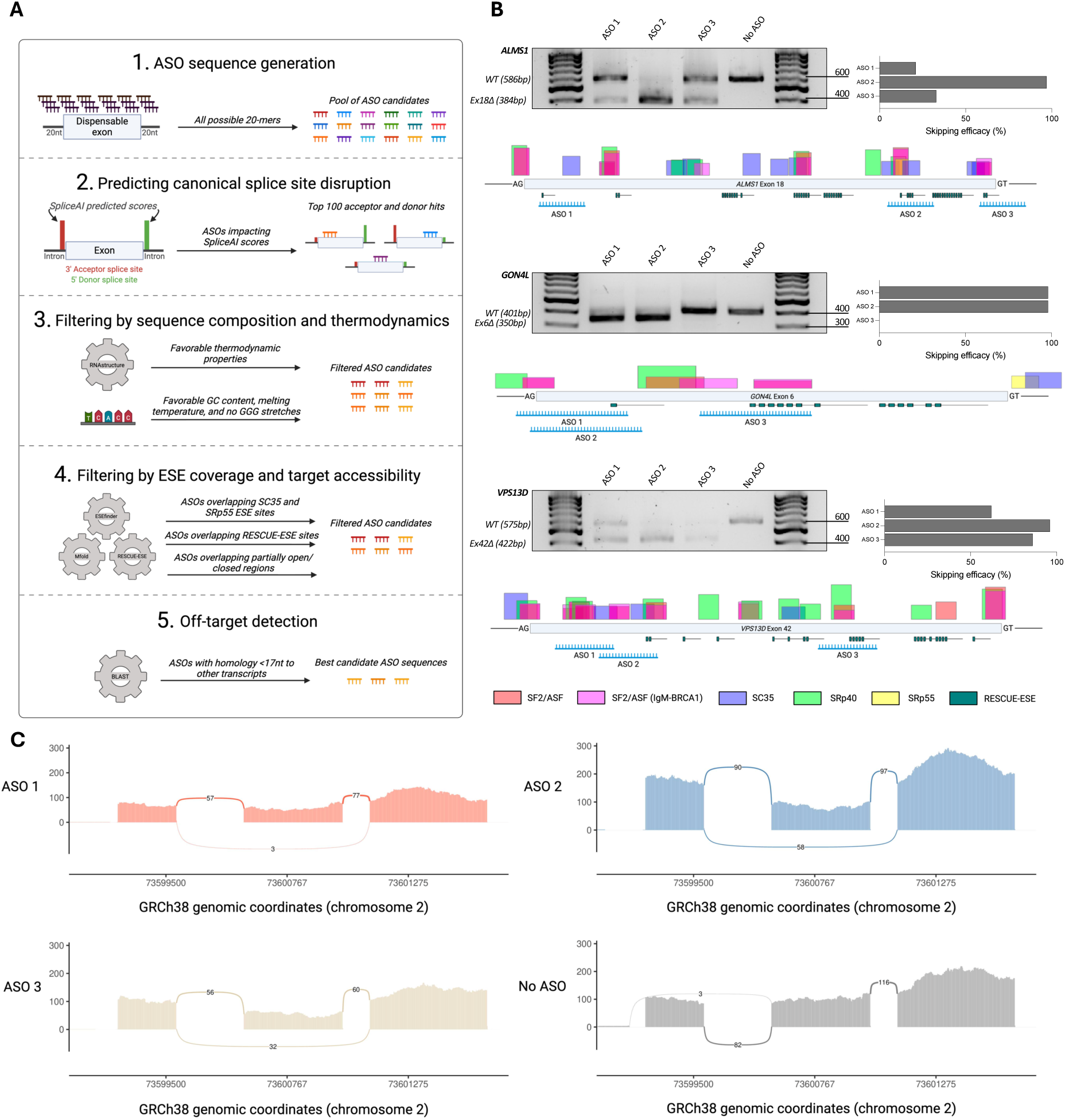
*In silico* design and functional validation of exon-skipping antisense oligonucleotides. (A) Schematic of *in silico* antisense oligonucleotide (ASO) design pipeline. All possible 20-mer ASO sequences spanning the target exon and into flanking introns are generated and ranked based on predicted disruption of canonical splicing. Candidates are filtered by sequence composition and thermodynamic properties (e.g. GC content and GGG stretches), predicted overlap with exonic splicing enhancers (ESEs) and target accessibility, and finally off-target homology against the transcriptome. Figure created using Biorender.com (B) Experimental validation of select *in silico*-optimized ASOs. Representative RT-PCR gels show wild-type (WT) and exon-skipped transcripts following treatment of healthy control fibroblasts (GM05659) with ASOs targeting NM_001378454.1(*ALMS1*) exon 18, NM_001282860.2(*GON4L*) exon 6, and NM_015378.4(*VPS13D*) exon 42. Bar graphs quantify exon skipping efficiency for each ASO based on densitometric analysis. For each exon, a schematic depicting splicing elements and ASO target sites is displayed (visualized using Alamut™ Visual Plus). (C) Sashimi plots showing splice junctions across *ALMS1* in ASO-treated healthy control fibroblasts. ASO treatment results in increased exons 17-19 junction reads, consistent with exon 18 skipping. Canonical splicing is displayed in an untreated control.

### Multi-modal functional validation of exon dispensability and ASO-mediated exon skipping for a rare genetic condition

Lamb-Shaffer syndrome (phenotype MIM# 616803) is an ultra-rare genetic condition associated with intellectual disability^44^. There are no disease-modifying therapies nor any publicly announced therapy development programs. Lamb-Shaffer syndrome is caused by haploinsufficiency of *SOX5*^44^. We evaluated the functional consequences of deleting NM_006940.6(*SOX5*) exon 4 (ΔEx4), which was classified as Dispensable and contains pathogenic stop-gain variants in ClinVar, and of deleting NM_006940.6(*SOX5*) exon 6 (ΔEx6), which was classified as Indeterminate-Possibly Dispensable (Figure 4A,B).

**Figure 4.**
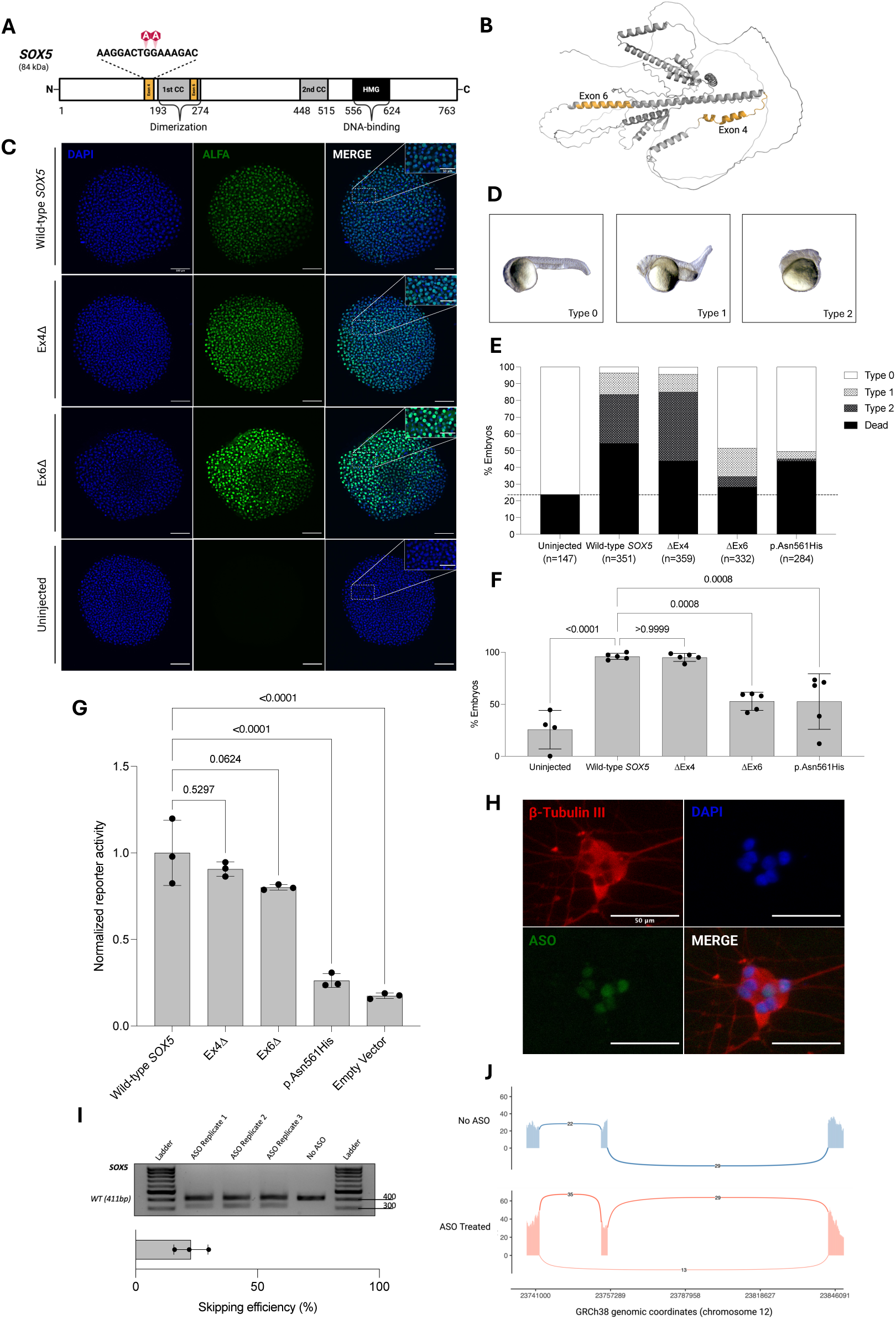
Functional validation of *SOX5* exon 4 and exon 6 skipping. (A) Schematic of the SOX5 protein. SOX5 contains two N-terminal coiled-coil domains mediating dimerization, and a C-terminal high-mobility group (HMG) box domain mediating DNA-binding. Positions of the target exons 4 and 6 are highlighted, as well as 2 pathogenic stop-gain variants reported in ClinVar. Figure created using Biorender.com (B) Predicted three-dimensional structure of SOX5 highlighting the locations of exons 4 and 6 within the protein. Structure was predicted using AlphaFold 3 and visualized using PyMOL. (C) Immunofluorescence analysis of SOX5 localization in zebrafish embryos at 6 hours post-injection of ALFA-tagged mRNAs for wild-type *SOX5*, ΔEx4, and ΔEx6. Nuclei were stained blue with DAPI, ALFA-SOX5 was stained green. Representative merged images illustrate nuclear localization across conditions. An uninjected embryo is shown as negative control. (D) Representative images illustrating scoring of zebrafish embryos at 24 hours post-injection. Type 0 (normal), Type 1 (mildly disrupted) and Type 2 (severely disrupted) are shown. (E) Distribution of developmental perturbation of zebrafish embryos at 24 hours post injection of wild-type *SOX5*, ΔEx4, ΔEx6, or p.Asn561His mRNAs. Stacked bars show the percentage of embryos classified as Type 0, Type 1, Type 2, or dead. Total numbers scored per condition are indicated. (F) Percentage of total disrupted embryos (Type 1, Type 2, or dead) across conditions. Wild-type *SOX5-* and ΔEx4-injected embryos show comparable disruption rates, whereas ΔEx6- and p.Asn561His-injected embryos exhibit significantly reduced disruption. Bars represent mean ± standard deviation (s.d.), with individual data points shown. One-way ANOVA with Dunnett’s multiple comparisons was used to compare each condition to wild-type *SOX5* mRNA; p-values are shown. (G) Luciferase reporter assay in HEK-293 cells assessing SOX5 transcriptional activity. Wild-type SOX5, ΔEx4, and ΔEx6 exhibited comparable activity, whereas p.Asn561His showed significantly reduced activity. An empty vector served as a negative control. Bars represent mean ± s.d., with individual replicates shown. One-way ANOVA with Dunnett’s multiple comparisons was used to compare each condition to wild-type SOX5; p-values are shown. (H) Immunofluorescence analysis in human induced neurons (iNeurons) following antisense oligonucleotide (ASO) treatment targeting *SOX5* exon 4. β-Tubulin III (red) stains neuronal cytoskeleton, nuclei are labeled with DAPI (blue), and ASO signal is green. A representative merged image is shown. (I) Evaluation of *SOX5* exon 4 skipping in iNeurons following ASO treatment. RT-PCR analysis shows shorter amplicon consistent with exon 4 skipping, with a mean efficiency of 22.7%. (J) Sashimi plots showing splice junctions across *SOX5* in ASO-treated iNeurons. ASO treatment results in increased exons 3-5 junction reads, consistent with exon 4 skipping. Canonical splicing is displayed in an untreated control.

To assess *in vivo* functional activity, we used a zebrafish model system. First, we assessed the ability of exon-skipped transcripts to localize in the typical way to the nucleus. mRNAs encoding N-terminally ALFA-tagged *SOX5* transcripts of interest were injected into 1-cell stage embryos. Protein localization was visualized using an anti-ALFA antibody at 6 hours post-injection. Wild-type SOX5, ΔEx4, and ΔEx6 were found to be exclusively localized to the nucleus, consistent with preserved subcellular localization (Figure 4C). Next, we performed a zebrafish overexpression assay comparing *SOX5* exon-skipped transcripts (ΔEx4 and ΔEx6, respectively) to wild-type and to a missense variant with known loss-of-function effects [NM_006940.6(*SOX5*):c.1681A>C p.(Asn561His)]^44^. Human *SOX5* mRNAs (wild-type, ΔEx4, ΔEx6 and p.Asn561His) were injected at the 1-cell stage, and embryos were evaluated for developmental perturbation phenotypes at 24 hours post-injection (Figure 4D,E). Overexpression of wild-type *SOX5* resulted in widespread developmental perturbation in 95.0% of embryos (Figure 4F). Embryos injected with ΔEx4 mRNA exhibited a similar degree of developmental perturbation to wild-type (96.0%), consistent with preserved *SOX5* activity (Figure 4F). In contrast, embryos injected with ΔEx6 or the pathogenic p.Asn561His variant showed significantly reduced developmental perturbation (52.8%, p=0.0008 compared with wild-type, and 52.5%, p = 0.0008 compared with wild-type, respectively) (Figure 4F).

To directly assess the transcriptional activity of ΔEx4 and ΔEx6 *in vitro*, we employed a luciferase assay in HEK293T cells using a reporter with an enhancer region synergistically activated by SOX5 and SOX9^44^. Co-expression of wild-type SOX5 and SOX9 resulted in a robust increase in luciferase reporter activity compared to an empty expression vector control (p < 0.0001) (Figure 4G). In contrast, expression of the known loss-of-function p.Asn561His variant resulted in significantly lower luciferase activity compared to wild-type (p < 0.0001) (Figure 4G). Expression of ΔEx4 enhanced SOX9-mediated activation of the luciferase reporter to a level comparable to wild-type (p = 0.5297) and significantly different from the pathogenic p.Asn561His variant (p < 0.0001) (Figure 4G). Expression of ΔEx6 enhanced SOX9-mediated activation of the luciferase reporter to a lesser but non-significantly different degree than wild-type (p = 0.0624), with results significantly different from the pathogenic p.Asn561His variant (p < 0.0001) (Figure 4G).

Overall, *in vivo* and *in vitro* assays provided empirical support for preserved SOX5 function after removing exon 4, and against preserved SOX5 function after removing exon 6. Last, we evaluated the feasibility of exon 4 skipping in a disease-relevant human cell model using an ASO. Human induced pluripotent stem cells were differentiated into induced neurons (iNeurons) and transfected with 100nM of an *in silico*-optimized exon-skipping ASO targeting *SOX5* exon 4 (Figure 4H). ASO effects were assessed at the transcript level using RNA extracted from iNeurons. RT-PCR followed by gel electrophoresis revealed a shorter amplicon consistent in size with exon 4 skipping. The exon-skipping efficiency of this ASO was quantified by densitometric analysis performed in triplicate, revealing a mean skipping efficiency of 22.7% (Figure 4I). Bulk RNA-sequencing of a single replicate revealed a skipping efficiency of 29%, consistent with densitometry-based quantification (Figure 4J).

## DISCUSSION

The continued growth of interventional genomics as a medical field is dependent on streamlining each of the complex steps involved in genetic therapy development, administration, and access^45,46^. Existing approaches to facilitate exon-skipping ASO development include curated databases of previously successful exon-skipping ASOs and tools for ASO design, with a primary focus being optimizing ASOs at the sequence level rather than exon dispensability^47–49^. We present the foundations for a scalable platform for exon-skipping ASO therapy development for inborn genetic diseases. HAWK-EYE is intended to address major upstream challenges in initiating ASO programs, by facilitating timely recognition of DNA variants potentially amenable to exon skipping as a therapeutic strategy, by sharing *in silico*-optimized ASO sequences for first-pass functional studies, and by recording results from experimental work confirming or refuting exon dispensability. Our findings from a Mendeliome-wide analysis highlight single exon skipping as an underrecognized and potentially undervalued therapeutic modality.

The prospect of an exon-skipping ASO therapy has been investigated for only a few diseases^22–37^. We piloted a gene-agnostic approach to (i) the identification of Dispensable exons, (ii) the design of exon-skipping ASOs, and (iii) the functional validation of exon dispensability. Although, and as expected, most exons are Indispensable, the HAWK-EYE database lists thousands of new candidate Dispensable exons in disease genes. Exon-skipping ASOs could be explored as a therapeutic intervention for individuals with P/LP variants in these Dispensable exons. Based on existing databases, such variants are continually being identified in clinical diagnostic laboratories around the world. In a diagnostic setting, HAWK-EYE may also aid in contextualizing the pathogenicity or benignity of whole-exon deletions. HAWK-EYE can support a shift from reactive to proactive identification of individuals with rare genetic diseases who could be nominated for ASO therapy development or, in a future state, receive an already developed and tested ASO.

We demonstrated a high experimental success rate for skipping for the *in silico*-optimized ASOs targeting Dispensable exons. Skipping efficiencies determined from RNA sequencing were lower than the densitometry-based estimates, consistent with the known amplification bias of RT-PCR toward shorter amplicons^15^; however, the relative ranking of ASO skipping efficiencies was preserved. By providing prioritized ASO sequences for each Dispensable exon, HAWK-EYE reduces barriers to initiating point-of-patient early-phase feasibility studies. Our open-access Mendeliome-wide database is particularly important for ultra-rare diseases, where commercial opportunities and industry partners are more limited. We emphasize that ASO sequences generated for the HAWK-EYE database are not suitable for immediate, direct translation to human clinical trials. Instead, high-throughput functional evaluation of dozens or hundreds of ASOs on patient-derived cell lines remains a cornerstone of the drug development process.

Robust functional evidence supporting exon (in)dispensability can lead to a change in primary category, but these data are rarely available, require manual review and curation, and were not incorporated into our genome-wide assessment. As proof-of-concept, we demonstrated how bespoke experiments feasible in many laboratories worldwide can be designed to follow-up on priority exons for skipping. The functional consequences of *SOX5* exon 4 deletion and of *SOX5* exon 6 deletion were compatible with our *in silico* dispensability classifications for these exons (Dispensable, and Indeterminate-Possibly Dispensable, respectively). Skipping exon 4 preserved *in vivo* nuclear localization and function, as well as *in vitro* transcriptional activity. Exon 4 is upstream of both N-terminal coiled-coil domains that mediate dimerization to enhance DNA binding, and of the C-terminal high-mobility group box that functions in site-specific DNA binding and protein localization^50^. Although exon 6 skipping preserved nuclear localization and, to a lesser extent, *in vitro* transcriptional activity, loss of function was observed *in vivo*. This discordance is likely because of limitations of the luciferase reporter assay, in which enhancers/promoters are overly accessible due to absent DNA methylation and chromatin architecture^51^. Exon 6 partially encodes the first coiled-coil domain of SOX5 that is known to be required for dimerization^52^. The classification of SOX5 exon 6 in the HAWK-EYE web application was subsequently updated to include an annotation based on this functional evidence, indicating that exon 6 is Indispensable. Similarly, clinical observations - in the form of DNA variants causing in-frame exon skips (e.g., canonical splice site variants^53^, full exon deletions) that are pathogenic or benign for disease – can also lead to a change in primary category. Requests to modify HAWK-EYE entries based on published or unpublished data germane to exon classifications can be submitted via the e-mail contact at https://hawk-eye.research.sickkids.ca/.

The accuracy of exon dispensability predictions will improve over time, with advances in multiplex functional assays and high throughput *in silico* protein modeling^54^. We relied on existing protein annotation resources and variant databases, which vary in completeness across genes and are biased towards extensively studied proteins. The HAWK-EYE database will be updated periodically to incorporate new data from any source. We focused on a single representative transcript (MANE Select), and incorporating isoform- and tissue-specific considerations is a future consideration. We recognize that exon- or sequence-specific constraints can limit ASO efficiency, and more generally that exon dispensability is necessary but not sufficient to consider an exon-skipping ASO as a realistic therapeutic intervention for an inborn genetic disease. Emerging technologies may be able to overcome exon- or sequence-specific constraints on ASO efficiency, such as next-generation base editing of acceptor and donor splice sites simultaneously^55^. Assessing disease amenability and delivery to target tissue(s) for each gene was out of scope, but may be addressed through other initiatives (N=1 Collaborative, personal communication). Clinical trials were also out of scope, with the expectation that our upstream efforts will accelerate exon-skipping ASO programs without limiting the potential for ASO patenting.

Customizable nucleic acid therapeutics - including ASOs and CRISPR-based gene editing - are nearing an inflection point. Proofs-of-concept for individualized genetic interventions, amid the challenges posed by extreme allelic heterogeneity and disease rarity, are leading to calls for platform approaches to development, regulated clinical trials, and approvals. The HAWK-EYE database is an attempt to shift the paradigm from reactive to proactive ASO design for ultra-rare variants and diseases and to enable rapid bespoke therapy development on disease-relevant timescales, beginning with exon skipping before expanding to allele-specific knock-down and splice-switching.

## METHODS

### Development of a high-throughput pipeline for predicting exon skippability

HAWK-EYE (**H**igh-throughput **A**nnotation **W**or**K**flow for dispensable **E**xon **Y**ield and **E**valuation) is a database that was developed using a custom pipeline implemented in R (v4.5.2)^56^. Additional functionality was provided by packages including *tidyverse*, *data.table*, *Biostrings*, *httr*, and *jsonlite*. HAWK-EYE integrates data from online databases and tools (Figure 1A), and generates exon-level annotations used to assess exons for eligibility towards canonical exon skipping in accordance with the N1C VARIANT guidelines version 1.0^18^. Exon coordinates and sequence information for Matched Annotation from NCBI and EMBL-EBI (MANE) Select transcripts are derived from RefSeq^57^ (NCBI Reference Sequence Database) and CCDS (Consensus Coding Sequence database)^58^. These data are used to determine exon length, cDNA size, exon frame, exon phase, and if a new codon is formed as a result of in-frame exon skipping. Exon position (including genomic coordinates in GRCh38) and sequence information from RefSeq are used to obtain variables from UniProt^59^, InterPro^60^, and ClinVar^61^. Specifically, the position of functional domains, compositional biases, repeat domains, motifs, zinc fingers, protein regions, and post-translational modifications was extracted from UniProt via its REST API (date accessed: December 2 2025), while predicted domain data were retrieved from InterPro via its API (date accessed: December 2 2025). The number of Pathogenic (P) or Likely Pathogenic (LP) missense, nonsense, and in-frame deletion variants per each exon was extracted from the ClinVar variant database (download date: December 2 2025).

### Defining a Mendeliome gene list for input into HAWK-EYE

We curated a list of human protein-coding genes associated with monogenic diseases, using three major online databases as input: Online Mendelian Inheritance in Man^62^ (OMIM; URL: https://www.omim.org/, download date: October 2025), ClinGen^63^ (URL: https://clinicalgenome.org/, download date: October 2025), and Genomics England PanelApp^64^ (URL: https://panelapp.genomicsengland.co.uk/, download date: October 2025). We excluded OMIM gene entries that only contribute to abnormal test values, to disease susceptibility, or that have a provisional understanding of disease association (denoted by symbols {}, [], and ?, respectively), retaining 4,318 genes with disease-gene associations. We excluded ClinGen gene entries with “limited”, “disputed”, or “refuted” supportive evidence classifications, entries with no known disease relationship, and entries with a mitochondrial DNA mode of inheritance, retaining 2,232 genes with disease-gene associations. We excluded PanelApp entries with “low” evidence, entries that listed regions/loci rather than specific genes, and entries with “unknown” or mitochondrial DNA mode of inheritance, retaining 3,437 genes with disease-gene associations. The union of these three sets was 5,098 unique genes (Supplementary Table 1); 41 genes were missing information (Supplementary Table 2) required for downstream annotations and filtering (see below), leaving 5,057 genes that we defined as the “Mendeliome” for our analyses. We obtained NCBI RefSeq accession numbers corresponding to MANE Select transcripts of the genome assembly GRCh38 for each gene. MANE Select transcripts were chosen to represent the most biologically relevant protein-coding sequences for each gene. The 5,057 genes comprised a total of 72,644 coding exons.

### Applying exon skipping eligibility criteria across the Mendeliome

Exon dispensability was assessed using an automated R-based workflow, and was informed by the primary considerations outlined in the N1C VARIANT guidelines version 1.0: position, frame context, and functional essentiality. We assigned all MANE Select exons in the Mendeliome to one of three primary categories: Indispensable, Indeterminate, and Dispensable. Specific *in silico* filtering parameters were optimized using sets of exons with functional evidence available in the published literature (Supplementary Table 3). Using the HAWK-EYE exon classification schema below, only 1/16 (6.2%) in-frame exons for which exon-skipping ASOs have been developed or proposed based on targeted experimental work were classified as Indispensable. Similarly, only 97/866 (11.2%) in-frame exons from Mendeliome essential genes identified by a large-scale CRISPR exon-level knockout screen as likely fitness-suppressing were classified as Dispensable^65^.

### Indispensable exons

Exons meeting any of the following criteria were classified as Indispensable:

i. part of the untranslated region;
ii. first or last coding exon;
iii. creates a new frameshift or premature termination codon if skipped;
iv. encodes the only annotated functional domain(s)/repeat(s) of the gene’s associated protein in UniProt;
v. encodes ≥10% of the coding sequence (CDS) and >1 functional domains/repeats in UniProt; or
vi. disease-causing (P/LP) complete exon, or nested in-frame, deletion reported in ClinVar.

### Dispensable exons

Exons satisfying all of the following criteria were classified as Dispensable:

i. not Indispensable;
ii. encodes <10% of the coding sequence (CDS);
iii. does not encode any annotated functional domain(s)/repeat(s) or other functional elements of the gene’s associated protein in UniProt, nor any post-translational modification sites or other amino acid-level elements, including residues such as glycosylation sites, active sites, and binding sites;
iv. does not encode any predicted functional elements in InterPro; and
v. does not contain any P/LP missense variants in ClinVar (as the presence of these types of variants suggests that the exon has a specific function, although no domain might be annotated or predicted).

### Indeterminate exons

Indeterminate exons were neither Indispensable nor Dispensable, and were further stratified into three mutually exclusive tiers (secondary categories) based on increasingly strict size and functional annotation considerations.

Unlikely Dispensable is the lowest confidence subset of Indeterminate exons, and includes exons meeting any of the following criteria:

i. encodes ≥10% of the CDS (regardless of annotated functional domain(s)/repeat(s)); or
ii. encodes ≥2/3 of the total number of any specific domain/repeat of the gene’s associated protein in UniProt.

Probably Dispensable is the highest confidence subset of Indeterminate exons, and includes exons meeting all of the following three criteria:

i. encodes <10% of the CDS;
ii. does not encode any functional domain/repeat or other functional elements (such as topological, transmembrane, and coiled-coil domains, regions, compositional biases, motifs, and modified residues [excluding lipids, glycans, and protein cross-links]);
iii. additional qualifiers as follows:

a. encodes any post-translational modification sites or other amino acid-level elements of the gene’s associated protein in UniProt, including residues such as glycosylation sites, active sites, and binding sites; OR
b. encodes any predicted functional elements in InterPro; OR
c. contains any P/LP missense variants in ClinVar.

Possibly Dispensable is the intermediate-confidence subset of Indeterminate exons, and includes exons meeting the following criteria:

i. neither Unlikely Dispensable nor Probably Dispensable;
ii. encodes <2/3 of the total number of any specific domain/repeat of the gene’s associated protein in UniProt; or
iii. encodes a functional element (such as topological, transmembrane, and coiled-coil domains, regions, compositional biases, motifs, and modified residues [excluding lipids, glycans, and protein cross-links]).

### Development of a high-throughput approach for exon-skipping ASO design

We developed a Python-based tool to generate and prioritize 20-mer exon-skipping ASO sequences. This tool accepts as input the sequence of a target exon and its flanking introns (±20 bp). All possible 20-mer ASO sequences are first generated by tiling across the input sequence and then ranked based on their predicted ability to perturb splice site recognition with SpliceAI^66^. Additional existing computational tools are incorporated into the pipeline to filter candidate ASO sequences in accordance with established exon-skipping ASO design principles^67^:

- ESEFinder^68^ and RESCUE-ESE^69^ are used to assess the potential of ASOs to disrupt exon recognition;
- RNAstructure^70^ and mFold^71^ are used to evaluate thermodynamic properties and target RNA secondary structures; and
- BLAST^72^ is used to assess potential off-targets against the human transcriptome.

The top ranked ASO sequences are filtered based on sequence composition and thermodynamic constraints, including %GC content, absence of ≥3 consecutive Gs (guanines), limited self-structure and dimerization energies, and favourable ASO-target binding energies. Target accessibility is evaluated using RNA secondary structure predictions, and ASO sequences are prioritized for targeting regions predicted to be accessible and likely to influence exon recognition. Off-target filtering excludes any sequences with ≥17 consecutive base pair matches to non-target sequences. ASO sequences failing any filtering criterion were excluded, and sequences satisfying all criteria were retained as *in silico*-optimized candidates. For use as experimental controls, corresponding scrambled ASOs can be created at https://www.genscript.com/tools/create-scrambled-sequence.

### Comparative analyses of exon classes

Exon inclusion levels were estimated using GTEx V10 (download date: December 2 2025) by calculating the proportion of transcripts across all tissues for a gene that contain each exon of interest. Nucleotide conservation scores were annotated using PhyloP^40^ and averaged by exon length to generate exon-level conservation scores (download date: December 2 2025). GC content was calculated directly from exon sequences as the proportion of guanine and cytosine bases per exon. For each metric, “low” scoring exons were defined as those at or below the 20^th^ percentile for all Mendeliome exons (Supplementary Table 4).

MANE Select transcript expression levels (TPM; Transcripts Per Million) were retrieved from GTEx for brain, liver, fibroblasts, and EBV-transformed lymphocytes. Since GTEx reports expression across multiple brain regions, transcript expression in the brain was assessed using the maximum TPM value observed across the 13 annotated brain tissues for each transcript. The EyeGEx^73^ dataset was retrieved from The Human Protein Atlas^74^ (URL: https://www.proteinatlas.org/, download date: December 2 2025), for retinal tissue expression levels. The threshold for detection was 0.5 TPM (Supplementary Table 5) Disease-associated (pathogenic or likely pathogenic) single-nucleotide variants (SNVs) and small insertions/deletions (indels; ≤ 50 bp) were extracted from public and private human genomic variation resources, including ClinVar (download date: May 6 2026)^75^, Genomics England (GEL release v19)^76^, Leiden Open Variation Database (LOVD v.3.0)^77^, and MSSNG^78^ (PMID: 36368308) (Supplementary Table 6). ClinVar variant data were retrieved from the publicly available variant_summary.txt file and filtered for germline variants with GRCh38 genomic locations in genes included in the Mendeliome. GEL variant data were retrieved from the Research Environment using the Exit Questionnaire (gmc_exit_questionnaire) table via the LabKey API, querying for genes included in the Mendeliome. LOVD variant data were retrieved using the REST API, querying for genes included in the Mendeliome. MSSNG variant data were retrieved from a supplementary dataset associated with the published study^78^. Unique variants across all resources were identified using HGVS genome annotation.

Gene-disease subgroup associations were obtained from GEL by downloading curated gene panels annotated with relevant Level 2 disease subgroups (download date: April 2026). For each disease subgroup, the corresponding gene set was compiled and intersected with genes containing Dispensable exons, genes containing Dispensable exons in which a disease-associated variant resides, as well as the Mendeliome as a whole. Overlap between disease subgroup-associated genes and genes containing Dispensable exons/Dispensable exons in which a disease-associated variant resides and the Mendeliome was assessed (Supplementary Table 7)

### Exon-skipping antisense oligonucleotide synthesis

All ASOs were synthesized by Integrated DNA Technologies (IDT) as 20-mers uniformly modified with 2’-O-methoxy-ethyl ribose and phosphorothioate linkages (Supplementary Table 8). Fluorescent ASOs were modified with a 5’ 6-carboxyfluorescein. Upon receipt, ASOs were resuspended in nuclease-free water to a stock concentration of 100µM.

### Cell culture and ASO transfection

Healthy control fibroblasts (GM05659) were obtained from Coriell Cell Repositories (Camden, USA) and maintained in Dulbecco’s Modified Eagle Medium (DMEM) supplemented with 10% fetal bovine serum (FBS). ASO transfections were performed using Lipofectamine 3000 (Thermo Scientific L3000008), with transfection medium prepared in Opti-MEM (Thermo Scientific 31985062). A final concentration of 100nM of each ASO was incubated on cells for 16 hours, after which media was replaced with fully supplemented DMEM. At 48 hours post-transfection, cells were trypsinized (Wisent Bioproducts, 325-043-CL) and pellets were collected for RNA analysis.

Wild-type induced pluripotent stem cells (iPSCs; SCTi004-A) were obtained from STEMCELL Technologies and maintained in mTeSR™ Plus Basal Medium supplemented with mTeSR™ Plus 5X Supplement (STEMCELL Technologies, 100-0276). Induced neurons (iNeurons) were generated using the STEMdiff™-TF Forebrain Induced Neuron Differentiation Kit (STEMCELL Technologies, 100-1678) according to the manufacturer’s instructions. At day 5 of differentiation, ASO transfections were performed using Lipofectamine 2000 (Thermo Scientific, 11668030), with transfection medium prepared in Opti-MEM. A final concentration of 100 nM of each ASO was incubated on cells for 16 hours, after which media was replaced with BrainPhys™ Neuronal Medium (STEMCELL Technologies, 05797) supplemented with STEMdiff™ Forebrain Neuron Maturation Supplement (STEMCELL Technologies, 08606). At 48 hours post-transfection cells were trypsinized and collected for RNA analysis or fixed for use in immunofluorescence microscopy.

### RNA isolation and reverse transcription polymerase chain reaction (RT-PCR) analysis

Cells were lysed, and total RNA was extracted using the Monarch® Spin RNA Isolation Kit (Mini) (NEB, T2110L). For fibroblast samples, 500ng total RNA was input for cDNA synthesis using the iScript™ cDNA Synthesis Kit (Bio-Rad, 1708890), whereas 100ng total RNA was input for iNeuron samples. 2µL of cDNA synthesis product was then used as input for a 25µL PCR reaction using Q5® High-Fidelity 2X Master Mix (NEB, M0492). Primer sequence, corresponding annealing temperatures and extension time are listed in Supplementary Table 9. PCR products were resolved on a 2% agarose gel prepared in 1x TAE buffer and imaged using the GelDoc™ Go Gel Imaging System (Bio-Rad; Hercules, CA, USA). Densitometric analysis of PCR bands was performed using ImageJ software (NIH) to quantify exon skipping by measuring relative intensities of skipped and wild-type amplicons.

### Bulk RNA-sequencing sample processing and analysis

Illumina Stranded poly(A) mRNA library prep kit (NEBNext) was used, and samples were run on Illumina NovaSeq X – 10B flowcell (Illumina, San Diego, CA, USA) using paired-end read. Raw RNA data was first trimmed with fastq (v0.23.4)^79^ and aligned to GRCh38 genome using STAR (v2.7.0f)^80^ in the two-pass mode. The aligned bams were visualized on IGV. Sashimi plots were created using R package ggsashimi^81^. Exon skipping efficiencies were calculated using splice-junction reads, where SJC denotes skipping junction counts, and UJC and DJC denote upstream and downstream inclusion junction counts respectively:

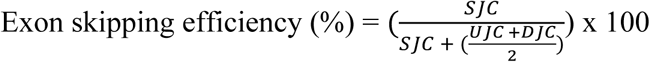

Analysis was performed at the High-Performance Computing Facility, Centre for Computational Medicine, The Hospital for Sick Children, Toronto, Canada.

### Immunofluorescence staining and imaging of iNeurons

At 48 hours post-ASO treatment, iNeurons were washed 3 x with room temperature Dulbecco’s Phosphate-Buffered Saline (DPBS). Cells were then fixed with 4% paraformaldehyde in phosphate-buffered saline (PBS) for 20 minutes at room temperature. PFA was removed, and cells were washed 3 x 5 minutes with PBS with rocking. Cells were then blocked in 0.2% Triton X-100, 3% sheep serum, and 1% Bovin Serum Albumin (BSA) in PBS for 1 hour at room temperature with rocking. Cells were then incubated overnight with mouse anti-beta-tubulin III antibody (1:1000; STEMCELL Technologies, 60052) in blocking solution at 4°C with rocking. Cells were washed 3 x 5 minutes with rocking and subsequently stained for goat anti-mouse Alexa Fluor 594 (1:500) and DAPI (1:1000) in PBS for 1 hour at room temperature with rocking. Cells were again washed with 3 x 5 minutes with PBS and imaged with an EVOS M5000 Imaging System (Thermo Fisher Scientific, Waltham, MA, USA).

### Plasmid construction, mutagenesis, and in vitro transcription

The full-length human *SOX5* coding sequence (NM_006940.6) was cloned into the pCSDEST vector (Addgene #22423) using Gateway LR cloning (Thermo Scientific, 12538120). Exon 4 and exon 6 deletions (*SOX5*ΔEx4, c.482_568del, and *SOX5*ΔEx6, c.742_810del), as well as a known loss-of-function missense variant (*SOX5* c.1681A>C, p.(Asn561His)), were introduced individually using the Q5 Site-Directed Mutagenesis Kit (NEB, E0552S). For immunofluorescence experiments only, an N-terminal ALFA-tag was added to wild-type and exon deletion *SOX5* constructs using site-directed mutagenesis. All constructs were sequence-verified by Sanger sequencing before use. Each plasmid was linearized in an overnight digest using NOTI-HF restriction enzyme (NEB, R3189L) and purified using the Monarch® Spin PCR & DNA Cleanup Kit (NEB, T1130). Linearized plasmids were *in vitro* transcribed into mRNA using the mMESSAGE mMACHINE SP6 kit (Thermo Scientific, AM1340), DNase-treated, and cleaned by column purification (NEB, T2040).

### mRNA microinjection

Wild-type *SOX5*, *SOX5*ΔEx4, *SOX5*ΔEx6, and *SOX5* p.Asn561His mRNAs were injected into AB zebrafish at the one-cell stage. Injections were performed using a pneumatic microinjection rig calibrated to a final injection volume of 1 nL. Microcapillary injection needles were prepared using a P-97 micropipette puller (Sutter Instrument, Novato, CA, USA). mRNAs were diluted in nuclease-free water and phenol red to a final injection concentration of 200 ng/µl and a final injection quantity of 200 pg of mRNA per embryo. Injection dosage was determined based on preliminary zebrafish experiments with wild-type *SOX5* mRNA.

### Embryo Phenotype Scoring

Injected embryos were maintained in E3 medium at 28.5°C and assessed at 24 hours post-injection for developmental abnormalities. A score of 0 was given to normal wild-type development, a score of 1 indicated mild developmental abnormalities, and a score of 2 indicated severe developmental arrest (Figure 4D). Embryos that were nonviable at the time of assessment were scored as dead. Embryo scoring was performed individually by two assessors, and scores were averaged for each condition. An uninjected control group of embryos was included in all experiments and evaluated using the same scoring criteria.

### Immunofluorescence staining and imaging of zebrafish embryos

Injected embryos maintained under standard conditions were collected at 6 hours post-injection and fixed in 4% PFA at 4°C overnight. Following fixation, embryos were washed with PBSTw (0.1% Tween-20 in PBS), dechorionated, and stored in 100% methanol until use. Embryos were rehydrated with a MeOH/PBSTw dilution series (100% MeOH, 75% MeOH/25%PBSTw, 50% MeOH/50%PBSTw, 25% MeOH/75%PBSTw), followed by 3 x 5 minutes PBSTw washes. Embryos were then washed in ice cold acetone at -20°C for 20 minutes, followed by a 5 minute wash in PBSTw and 3 x 10 minutes washes in PBSTr (1% Triton X-100 in PBS). Embryos were then blocked in 10% FBS and 5% BSA in PBSTr for 1 hour at room temperature. Embryos were then incubated overnight with mouse anti-ALFA antibody (1:250; NanoTag N1582) in blocking solution at 4°C. Embryos were washed 8 x 20 minutes in PBSTr and then stained for goat anti-mouse Alexa Fluor 488 (1:500) and DAPI (1:1000) in blocking solution at 4°C overnight. Embryos were again washed 8 x 20 minutes in PBSTr, followed by 2 x 5 minutes wash in PBSTw. Embryos were mounted with low-melting-point agarose in glass bottom imaging dishes and imaged with a Nikon AR1 Confocal Microscope (Nikon Instruments, Tokyo, Japan).

### Zebrafish lines and husbandry

Adult wild-type AB strain zebrafish were raised and maintained at 28.5°C in the zebrafish facility at the Hospital for Sick Children. Embryos were produced by natural mating of AB adults. All experiments were approved by the Animal Care Committee at the Peter Gilgan Centre for Research and Learning and were performed strictly in accordance with the Guidelines of Canadian Council on Animal Care.

### Luciferase reporter assay

The [4xA1]-p89Luc reporter plasmid and expression plasmids encoding SOX9 and SOX5 were generated as previously described and kindly provided by the authors^44^. *SOX5* exon 4 and 6 deletions, and loss-of-function missense variant *SOX5* c.1681A>C, were generated by site-directed mutagenesis of the wild-type *SOX5* plasmid (Q5 Site-Directed Mutagenesis Kit; NEB, E0552S). HEK-293 cells were co-transfected with 50ng [4xA1]-p89Luc reporter, 5ng *SOX9* expression plasmid, 1ng pNL1.1.TK[Nluc/TK] (Promega, N1501), and 15ng of wild-type/exon deletion/variant *SOX5* expression plasmid using Lipofectamine 3000 (Thermo Scientific L3000008). At forty hours post-transfection, luminescence was measured with Nano-Glo® Dual Luciferase® Reporter Assay System (Promega, N1610) using the Varioskan™ LUX Multimode Microplate Reader (Thermo Fisher Scientific, Waltham, MA, USA). Reporter activities were calculated as means with standard deviation (s.d.) of luciferase values measured in triplicate and normalized for transfection efficiency using NanoLuc values.

### Protein modelling

The SOX5 protein structure was predicted using AlphaFold 3, with the human PDB sequence (NP_008871.3) obtained from the National Center for Biotechnology Information database. Structures were visualized using PyMOL (v3.1.3)^82^.

### Statistical methods

Statistical analyses were performed using GraphPad Prism 10. Data are presented as mean ± s.d. For luciferase and zebrafish overexpression assays, comparisons between groups were performed using one-way analysis of variance (ANOVA) followed by Dunnett’s multiple comparison test. Odds ratio with 95% confidence intervals were calculated for enrichment analyses. Sample sizes and p values are reported in corresponding figure captures. A two-sided p value < 0.05 was considered statistically significant.

## Data Availability

All HAWK-EYE classifications and ASO sequences are available through the HAWK-EYE web application. All scripts used to generate exon annotations and classifications of the HAWK-EYE database are available on Zenodo and on the GitHub repository.
Data from the National Genomic Research Library (NGRL) used in this research are available within the secure Genomics England Research Environment. Access to NGRL data is restricted to adhere to consent requirements and protect participant privacy. Data used in this research include: variant-level data from the Rare Disease programme Exit Questionnaire (gmc_exit_questionnaire) table, accessed via the LabKey API and filtered for clinically reported pathogenic and likely pathogenic variants associated with fully or partially solved cases.
Access to NGRL data is provided to approved researchers who are members of the Genomics England Research Network, subject to institutional access agreements and research project approval under participant-led governance. For more information on data access, visit:
https://www.genomicsengland.co.uk/research

https://hawk-eye.research.sickkids.ca/

https://zenodo.org/records/20560127

https://github.com/CostainLab/hawk-eye-exon-annotations

## ETHICS DECLARATION

No human participants or data were involved in this study. All animal experiments were approved by the Animal Care Committee at the Peter Gilgan Centre for Research and Learning and were performed strictly in accordance with the Guidelines of Canadian Council on Animal Care.

## AVAILABILITY OF DATA AND MATERIALS

All HAWK-EYE classifications and ASO sequences are available through the HAWK-EYE web application. All scripts used to generate exon annotations and classifications of the HAWK-EYE database are available on Zenodo^83^ and on the GitHub repository^84^.

Data from the National Genomic Research Library (NGRL) used in this research are available within the secure Genomics England Research Environment. Access to NGRL data is restricted to adhere to consent requirements and protect participant privacy. Data used in this research include: variant-level data from the Rare Disease programme Exit Questionnaire (gmc_exit_questionnaire) table, accessed via the LabKey API and filtered for clinically reported pathogenic and likely pathogenic variants associated with fully or partially solved cases.

Access to NGRL data is provided to approved researchers who are members of the Genomics England Research Network, subject to institutional access agreements and research project approval under participant-led governance. For more information on data access, visit: https://www.genomicsengland.co.uk/research

## COMPETING INTERESTS

The authors declare no competing interests.

## FINANCIAL DISCLOSURES

LN was supported by a Canada Graduate Scholarship-Master’s award from the Canadian Institutes of Health Research, an Ontario Graduate Scholarship, and a SickKids Restracomp Doctoral Scholarship. Development of the HAWK-EYE web application was supported by the Data Sciences Institute at the University of Toronto via grant number DSI-RSDY3R1P10. Experimental work was supported by a Rapid Impact Mini Grant from the Program in Genetics & Genome Biology (SickKids Research Institute), and the Azrieli Precision Child Health Platform. ARD is supported by the Canadian Institute for Health Research, PSI foundation, National Sciences and Engineering Research Council, SickKids Research Institute and the Azrieli Precision Child Health Platform.

## ACKNOWLEDGEMENTS

We thank members of the Translational Genomics and Advanced Therapeutics teams within SickKids Precision Child Health, and colleagues affiliated with the N=1 Collaborative, for thoughtful discussions related to this work. We thank Dr. Véronique Lefebvre for sharing plasmids used in this study. This research was enabled in part by support provide Compute Ontario (computeontario.ca) and the Digital Research Alliance of Canada (alliancecan.ca)

Data used for analyses described in this manuscript were obtained from the GTEx Portal on 12/02/2025.

We gratefully acknowledge the participates of the National Genomic Research Library (NGRL), whose contributions made this research possible. Secure access to the NGRL under project ID 1126 was provided by Genomics England, which delivers the NGRL in partnership with NHS England, and is wholly owned by the UK Department of Health and Social Care. The NGRL contains participants’ health data collected by the NHS as part of their care, along with samples and data from their participation in research, for which fully informed consent has been obtained. This includes genomic and clinical data provided through the NHS Genomic Medicine Service, as well as data obtained through research studies, including the 100,000 Genomes Project and the Generation Study, both of which are delivered in partnership with the NHS, and from other research cohorts involving external collaborators.

## AUTHOR CONTRIBUTIONS

Conceptualization: GC

Data curation: LN, BH, DC, CTT, CK, TQ, RMM, EAI

Formal analysis: LN, BH, RS, YL

Funding acquisition: GC, ARD

Supervision: GC, ARD

Visualization: LN, BH, LV, YL

Writing-original draft: LN, GC

Writing-review & editing: BH, DC, CTT, CK, RS, TQ, LV, YL, RMM, EAI, ARD

## Notes

### Competing Interest Statement

The authors have declared no competing interest.

